# Life after patent: Drug price dynamics and cost-effectiveness analysis

**DOI:** 10.1101/2023.06.08.23290510

**Authors:** Miquel Serra-Burriel, Nicolau Martin-Bassols, Gellért Perényi, Kerstin N. Vokinger

## Abstract

This study analyzed the effects of patent expiration on drug prices in eight countries (US, UK, Canada, Australia, Japan, France, Germany, and Switzerland) and the impact of these price dynamics in cost-effectiveness assessments. First, using an event study design, we showed that average prices of drugs substantially decreased eight years after patent expiration. Then, to assess the implications of this finding for cost-effectiveness assessments, a theoretical cost-effectiveness model simulated two real-world scenarios: (1) the comparator drug was a generic and the patent of the new drug expired after market entry; (2) the comparator drug was also under patent protection, but the patent expired prior to the patent of the new drug. Not accounting for genericization or patent expiration of the comparator drug resulted in an underestimation or overestimation of the incremental cost-effectiveness ratios, respectively. Our pricing dynamic estimates can be applied to base-case analyses of cost-effectiveness models.

## 1. Introduction

The loss of patent protection for originator drugs and biologics (for simplicity both are referred to as “drugs”) is a crucial moment for manufacturers, patients and society at large. After a period of profit internalization by the innovator, society can benefit from the innovation at lower prices due to the market entry of generic versions of the originator drug that results in price competition (Frank & Salkever, 1997). However, the level of competition after generics and biosimilars (follow-on class of biologics, for simplicity both are referred to as “generics”) enter the market varies strongly across markets and nations (Beall et al., 2020; Carl et al., 2022). Companies of originator drugs have an incentive to delay or preclude the genericization of their products to extend their temporary monopoly with different strategies resulting in antitrust lawsuits (Böhme et al., 2021; Vokinger et al., 2017).

Cost-effectiveness analyses (CEA) of new drugs and biologics are usually conducted as a basis for price negotiations and reimbursement decisions, and to improve the efficiency of healthcare systems (Clement et al., 2009). CEAs are based on comparative analyses – the costs and effectiveness of the new originator drug are compared to those of an already marketed drug with the same indication. Therefore, the price of therapeutic alternatives tends to be the most significant contributing factor when determining the cost-effectiveness profiles of drugs. In general, originator drugs are under patent protection at market entry and patent expiration follows sometime after market entry. Most CEA’s use a fixed, non-dynamic price for the therapeutic alternatives. In previous studies, the assumption that prices will remain constant has been criticized with the reasoning that this does not adequately reflect the real-world setting (Guertin et al., 2015), and there have been debates on whether price dynamics of drugs should be incorporated into CEA’s (Hirst et al., 2015; Schans et al., 2020). Some argued that CEA estimates are rendered biased by not incorporating future price dynamics (Shih et al., 2016). In contrast, others highlighted that the CEA estimates are contingent to the target population and the timing of the investment (Guertin et al., 2015). Despite all these discussions, there is, in any case, i) a lack of estimates of pricing dynamics before and after patent loss of the new originator drug or comparative drug, and ii) unclarity of their influence on cost-effectiveness estimates (Hoyle, 2008; Hua et al., 2019). A recent review assessed to which degree there is clear guidance on this issue, concluding that the omission of assumptions regarding genericization misguides long run opportunity costs assessments, and highlighting the need for further work in the area (Neumann et al., 2022).

The two most common real-world scenarios for the CEA of new originator drugs involve a comparison with either a generic drug or another originator drug under patent. In the first scenario, the price of the new originator drug is expected to substantially decrease after patent expiration, i.e., to improve its cost-effectiveness profile over time. As a result, its incremental cost-effectiveness ratio is presumably underestimated. In the second scenario, the patent of the comparator drug is expected to expire earlier than the patent of the new originator drug. In this case, ignoring the pricing dynamics will likely result in an overestimation of the cost-effectiveness of the new originator drug by assuming that the price of the comparator stays constant, and, thus, estimate a higher than the actual price after patent expiration.

In this study, we present for the first time a comprehensive genericization pricing dynamics estimation based on an event study design. First, we compared prices of new originator drugs before and after patent loss with drugs that did not experience patent loss in the same time period in eight major developed economies. Then, through simulation studies, we assessed the effects of incorporating such estimates into cost-effectiveness models based on their clinical profile and time to patent expiration. Overall, we found a steep progressive decrease in drug prices after patent expiration that varied significantly across countries. Finally, we created a theoretical cost-effectiveness model two simulate two common real-world scenarios: In the first scenario the new originator drug was compared with a generic drug, and the patent of the new originator drug expires after market entry. In the second scenario, both drugs – the new originator drug and the comparative drug – were under patent at the initial time point but the comparative drug lost patent prior to the new originator drug. For both scenarios we then analyzed how the incorporation of pricing dynamics affected the cost-effectiveness estimates. Our results demonstrate that in the first scenario the cost-effectiveness profile of the new originator drug is slightly underestimated, while in the second scenario the cost-effectiveness of the new originator drug is strongly overestimated.

The structure of this study is as follows: In section 2, we present both the statistical methods and data for the empirical analysis as well as the theoretical cost-effectiveness simulation model. In section 3, we outline the empirical and simulation results, and we discuss the results in section 4.

## 2. Methods

This study comprises two distinct parts: First, an empirical analysis of drug pricing dynamics, including the impact of price competition on drug prices before and after patent expiration, in eight countries (US, UK, Japan, Australia, Canada, France, Germany, Switzerland). Second, a theoretical cost-effectiveness model was constructed to simulate two real-world scenarios with incorporated pricing estimates.

### 2.1 Data

We use drug pricing data from IQVIA data (IMS health) for the time period 2011 to 2020 for eight countries –US, UK, Japan, Australia, Canada, France, Germany, Switzerland. The dataset contains quarterly information on list prices, units, companies, molecules, doses, and patent status for privately sold drugs. For the included countries, both retail and in-hospital sales data were available. Our unit of observation was at molecule level and our outcome was the unit-weighted average price for a given quarter. Table S1 presents the descriptive statistics of the analytical sample by included country, and Table S2 displays the observation availability with respect to the timing of the patent expiration alongside the patent status.

In our analytical sample, we included all drugs that were under patent protection at the start of the observation window. Drugs that lost their patent at any time point between 2011 and 2020 were classified in the “treatment” group, and those drugs that did not lose their patent in this time period were classified in the “control” group. Our data contained list prices adjusted for national inflation; we were not able to account for rebates (Kakani et al., 2020).

### 2.2 Empirical strategy

To identify the effects of patent expiration on drug prices, we leveraged the timing of patent expiration within countries between 2011 and 2020. We used quasi-experimental variation created by the staggered loss of patents, which under a certain set of assumptions allowed us to approximate the causal effect of patent expiration on drug prices with a difference-in-differences strategy. We estimated one model per country, independently.

The main assumption of our empirical strategy was that if drugs did not experience patent loss, their prices would have evolved in parallel to those under patent protection. Under this assumption, our model rules out that results are driven by constant price differences across drugs and common national pricing trends. To provide empirical support for this assumption, we estimated a dynamic version to examine the existence of pre-trends with the Callaway and Sant’Anna difference-in-differences estimator (Callaway & Sant’Anna, 2021). Another reason to use the estimator instead of a standard two-way fixed-effects model is that should the treatment effects be heterogeneous across time, the estimator would be inconsistent. The target estimand was the average treatment effect on the treated group (ATT), i.e., the effect of losing patent for those drugs for which the patent expired. Standard errors of the coefficients of interest were clustered at molecule level. We also performed the same model with number of competitors selling the molecule in the market as an outcome.

Finally, to provide suggestive evidence on the effect of competitor entry on price competition, we estimated the effect of one additional competitor in the market on the log price, using a panel design with country, substance, and time fixed-effects:

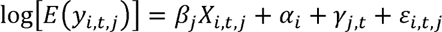

Where, y_i,t,j_ is the unit-weighted price of molecule *i*, at calendar quarter *t,* in country *j.* The coefficient of interest is {3_j_, the marginal effect of one additional competitor on the log price per country, and X_i,t,j_ is an indicator of the number of competitors selling a given molecule at a given time period in a given country. a_i_ represents molecule fixed-effects, and y_j,t_ country-quarter fixed-effects. Finally, c:_i,t,j_ is an idiosyncratic error term. We also included an additional specification with both linear and quadratic terms of number of competitors to provide suggestive evidence on the effect of the intensity of competition on prices.

We estimated the model with a quasipoisson specification, clustered the standard errors at the molecule-country level, and scaled the coefficients by two standard deviations (Gelman, 2008) to enhance comparability across countries. The estimates of this model must be interpreted cautiously as they represent a mechanistic approximation, and there might be factors not captured in the fixed-effects structure that simultaneously affect the price and the entry decision by competitors.

### 2.3 Theoretical cost-effectiveness modelling and simulation studies

To assess the influence of price dynamics after patent expiration on cost-effectiveness estimates, we created a simple health economic model. It compares two alternative treatments with survival of patients as the effectiveness metric. We created two scenarios that cover most scenarios found in health economic evaluations that aim to serve as a basis for price negotiation and reimbursement decisions.

In the first scenario, the originator drug was subject to patent protection for a time period prior to patent expiration, while the comparator drug was a generic. In the second scenario, both drugs, the originator and the comparator drug were under patent protection, with the latter losing patent protection prior to the originator drug.

The modelling was motivated by the fact that most economic evaluations of drugs occur when the new originator drug enters the market. In most cases, the new drug is under patent protection at market entry but will lose its patent within the modelling time window. The model includes incremental costs and survival effects discounted at a constant rate over the time horizon:

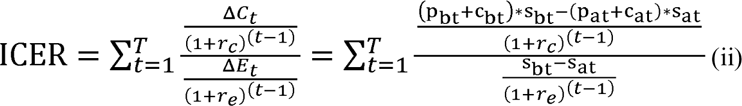

Where incremental effectiveness ΔE_t_ expressed as the difference in survival rates s_bt_ - s_at_ between intervention *b* and *a,* at time point *t.* Incremental costs ΔC_t_ are defined as the difference in drug price p_t_ plus non-drug costs c_t_ times the proportion of patients that survived S_t_ (assuming that patients receive therapy while alive) between interventions *b* and *a.* Both incremental costs and survival are discounted at r_e_ and r_e_ respectively. Derivations and special cases of the model are presented in the supplementary material.

We then incorporated the pricing estimates from the empirical assessment two, eight, and 14 years after target intervention introduction times. We presented three effectiveness scenarios with a different survival effects hazard ratios of (0.44, 0.58, 0.89), representing high, mid, and low effectiveness. We then assessed the extent of the influence that the introduction of pricing dynamics has depending on the abovementioned scenarios.

## 3. Results

### 3.1 Price levels

Overall, when comparing all eight included countries in the study, the US presented the highest drug prices, both on average and median terms. Mean list prices in the US were between 3.5 (Japan) and 1.6 (Germany) times higher for new originator drugs under patent protection, and 4.1 (Australia) and 1.7 (Switzerland) times higher for drugs without patent protection. The pattern was similar for median prices.

In terms of price developments over time, the US experienced the highest mean price increase over the 10-year period, with a 119% growth, while for the other countries the increase ranged from 10 to 30%. Table 1 provides the summary statistics of prices.

**Table 1.** Absolute mean and median drug price levels by country, year, and patent status in US dollars (2020).

| Year | Patent | Australia |  | Canada |  | France |  | Germany |  | Japan |  | Switzerland |  | UK |  | US |  |
| --- | --- | --- | --- | --- | --- | --- | --- | --- | --- | --- | --- | --- | --- | --- | --- | --- | --- |
|  |  | Mean | Median | Mean | Median | Mean | Median | Mean | Median | Mean | Median | Mean | Median | Mean | Median | Mean | Median |
| 2011 | Protected | 1254.7 | 680.3 | 2446.6 | 1010.4 | 2302.3 | 61.4 | 2399.3 | 536.6 | 1293.4 | 941.1 | 1969.4 | 15.8 | 1844.5 | 873.8 | 2556.3 | 589.6 |
| 2011 | Unprotected | 536.4 | 110.7 | 738.0 | 173.3 | 599.3 | 60.1 | 845.5 | 135.5 | 576.3 | 249.7 | 963.2 | 192.4 | 587.4 | 128.5 | 1133.4 | 354.2 |
| 2012 | Protected | 1256.9 | 603.2 | 2721.8 | 1459.5 | 1478.7 | 66.9 | 2930.4 | 1031.4 | 1482.6 | 865.1 | 2182.9 | 132.5 | 2410.3 | 867.7 | 3185.4 | 906.3 |
| 2012 | Unprotected | 585.9 | 117.3 | 840.9 | 184.1 | 586.6 | 57.5 | 851.4 | 132.8 | 552.1 | 242.0 | 996.1 | 184.1 | 605.4 | 181.6 | 1267.1 | 341.0 |
| 2013 | Protected | 1300.9 | 568.2 | 2634.7 | 1331.3 | 1342.0 | 150.5 | 3180.3 | 1265.9 | 1116.0 | 662.4 | 2161.5 | 150.0 | 2299.0 | 839.3 | 4031.2 | 975.5 |
| 2013 | Unprotected | 525.3 | 88.1 | 828.1 | 174.2 | 570.6 | 60.8 | 931.9 | 159.5 | 440.9 | 193.2 | 970.3 | 181.9 | 691.4 | 189.9 | 1361.7 | 353.6 |
| 2014 | Protected | 1535.1 | 620.4 | 2898.3 | 1361.6 | 1310.8 | 315.5 | 3014.0 | 1217.9 | 1068.0 | 707.6 | 2444.8 | 158.4 | 2621.7 | 904.4 | 4724.0 | 1526.5 |
| 2014 | Unprotected | 454.2 | 69.8 | 787.9 | 172.9 | 610.8 | 61.2 | 972.8 | 140.1 | 427.0 | 167.4 | 999.8 | 188.1 | 743.8 | 189.2 | 1392.7 | 382.8 |
| 2015 | Protected | 1319.0 | 380.8 | 2717.0 | 1186.8 | 1330.0 | 347.3 | 2828.9 | 1300.4 | 1235.0 | 644.0 | 2576.5 | 692.5 | 2533.6 | 1177.1 | 4745.5 | 1253.3 |
| 2015 | Unprotected | 339.1 | 51.8 | 676.3 | 148.0 | 490.3 | 50.1 | 984.6 | 121.1 | 412.9 | 147.5 | 1078.2 | 186.2 | 726.3 | 181.1 | 1734.8 | 463.3 |
| 2016 | Protected | 1466.3 | 391.8 | 2576.6 | 1092.7 | 1359.0 | 260.6 | 2937.4 | 1290.8 | 1369.9 | 714.5 | 2375.1 | 1039.6 | 2208.6 | 1075.3 | 4808.1 | 1439.5 |
| 2016 | Unprotected | 334.9 | 49.3 | 654.9 | 132.6 | 513.6 | 57.3 | 881.9 | 120.9 | 489.0 | 148.4 | 1038.3 | 204.8 | 726.0 | 149.8 | 1902.0 | 530.3 |
| 2017 | Protected | 1697.7 | 534.1 | 2720.6 | 1058.3 | 1385.8 | 587.1 | 2939.9 | 1428.8 | 1149.4 | 725.9 | 2634.1 | 1268.2 | 2243.8 | 1021.7 | 5089.4 | 1737.9 |
| 2017 | Unprotected | 354.1 | 48.9 | 663.0 | 129.0 | 597.9 | 62.6 | 861.0 | 132.5 | 506.4 | 137.5 | 1032.9 | 207.1 | 730.6 | 122.0 | 2097.2 | 519.7 |
| 2018 | Protected | 1634.7 | 475.4 | 2709.0 | 1073.0 | 2162.7 | 876.9 | 2973.9 | 1485.6 | 1273.9 | 806.2 | 2464.7 | 1180.3 | 2408.6 | 1060.3 | 5096.8 | 2040.7 |
| 2018 | Unprotected | 339.7 | 45.1 | 708.4 | 128.5 | 625.5 | 59.5 | 923.1 | 125.2 | 507.7 | 132.9 | 939.4 | 176.7 | 786.8 | 141.1 | 2103.0 | 431.7 |
| 2019 | Protected | 1536.0 | 440.1 | 2417.6 | 939.4 | 2137.0 | 907.8 | 2718.1 | 1324.0 | 1413.5 | 843.6 | 2219.9 | 1045.9 | 2352.3 | 1002.0 | 5395.2 | 2433.1 |
| 2019 | Unprotected | 338.8 | 39.9 | 694.3 | 118.1 | 595.0 | 56.9 | 883.7 | 115.8 | 493.4 | 123.7 | 883.2 | 161.3 | 803.8 | 127.7 | 1997.6 | 397.6 |
| 2020 | Protected | 1666.3 | 456.9 | 2588.2 | 1001.1 | 2227.4 | 899.1 | 2936.4 | 1413.3 | 1418.4 | 871.2 | 2435.3 | 1122.2 | 2434.1 | 1044.5 | 5586.8 | 2541.0 |
| 2020 | Unprotected | 315.3 | 35.2 | 690.5 | 120.1 | 571.7 | 57.4 | 833.4 | 116.2 | 492.2 | 119.6 | 922.8 | 162.0 | 817.7 | 121.4 | 1776.9 | 387.0 |

*Notes: control group: never treated, 0 anticipation periods, doubly robust estimation, Callaway-Sant'Anna staggered difference-in-differences method.*

### 3.2 Event study

Figure 1 presents the dynamic ATT estimates from our primary specification by country, and shows that after patent expiration, drug prices decreased in all countries. Our estimates pre-intervention also provide strong support for parallel trends in all countries, up to eight year pre-patent loss or 32 quarters.

**Figure 1.**
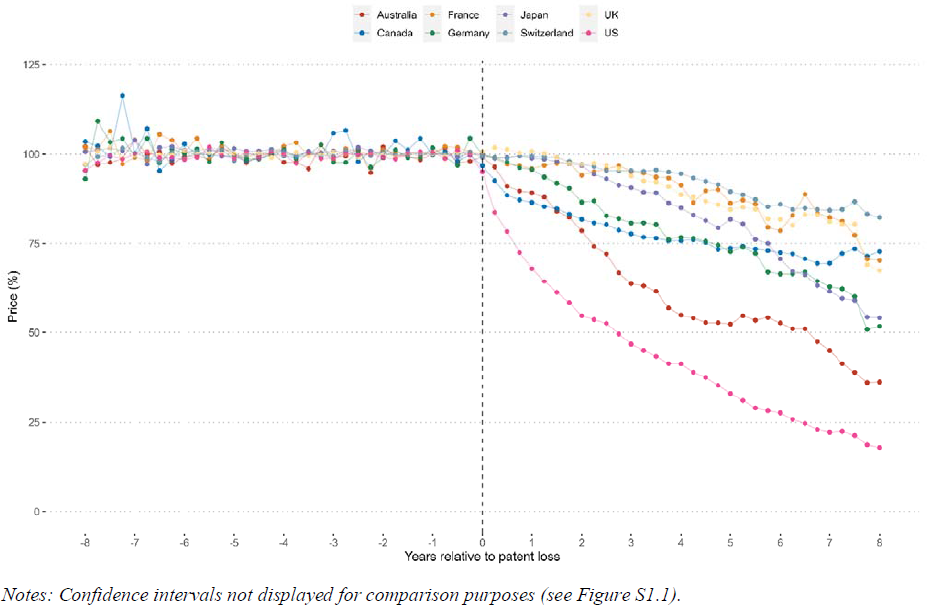
Difference-in-differences dynamics estimates by country.

Stark differences across countries were observed in terms of effect sizes. The US and Australia presented the steepest declines in prices, reaching 82% and 64% eight years after patent expiration in the US and Australia, respectively. Japan and Germany followed with an approximately 50% price decrease. Canada, France, Germany, and Switzerland presented the smallest price decrease with approximately 25%. Table 2 presents the numeric estimates and confidence intervals for each country, and Figure S1.1. presents all estimates of the countries with their respective confidence intervals.

**Table 2.** Annual drug price change estimates with respect to the period previous to patent protection expiration.

|  | Australia |  | Canada |  | France |  | Germany |  | Japan |  | Switzerland |  | UK |  | US |  |
| --- | --- | --- | --- | --- | --- | --- | --- | --- | --- | --- | --- | --- | --- | --- | --- | --- |
| T | Exp(L) | 95%CI | Exp(L) | 95%CI | Exp(L) | 95%CI | Exp(L) | 95%CI | Exp(L) | 95%CI | Exp(L) | 95%CI | Exp(L) | 95%CI | Exp(L) | 95%CI |
| -8 | 1.02 | 0.96-1.08 | 1.03 | 0.93-1.15 | 1.02 | 0.97-1.07 | 0.93 | 0.75-1.15 | 1.01 | 0.99-1.03 | 0.97 | 0.88-1.08 | 0.97 | 0.88-1.08 | 0.95 | 0.89-1.02 |
| -7 | 1.04 | 0.95-1.14 | 1.00 | 0.98-1.02 | 0.99 | 0.95-1.04 | 0.99 | 0.94-1.05 | 1.04 | 0.93-1.16 | 1.00 | 0.98-1.03 | 1.00 | 0.97-1.02 | 1.00 | 0.96-1.04 |
| -6 | 0.99 | 0.95-1.03 | 1.03 | 0.96-1.1 | 1.00 | 0.98-1.03 | 1.00 | 0.95-1.05 | 1.01 | 0.99-1.04 | 0.99 | 0.97-1.01 | 1.01 | 0.99-1.03 | 0.99 | 0.96-1.01 |
| -5 | 1.00 | 0.97-1.03 | 0.99 | 0.96-1.03 | 0.99 | 0.96-1.02 | 1.00 | 0.97-1.04 | 1.02 | 0.99-1.04 | 0.98 | 0.95-1.01 | 1.00 | 0.97-1.03 | 0.99 | 0.97-1.01 |
| -4 | 0.98 | 0.94-1.02 | 1.00 | 0.98-1.01 | 1.02 | 0.99-1.05 | 1.01 | 0.97-1.05 | 1.01 | 0.98-1.04 | 1.00 | 0.99-1.01 | 1.00 | 0.99-1.01 | 1.00 | 0.98-1.02 |
| -3 | 0.99 | 0.96-1.02 | 1.06 | 0.98-1.14 | 0.98 | 0.96-1 | 0.98 | 0.92-1.04 | 1.01 | 0.98-1.04 | 0.99 | 0.98-1.01 | 1.01 | 0.99-1.02 | 0.99 | 0.97-1.01 |
| -2 | 1.02 | 0.99-1.05 | 1.00 | 0.98-1.01 | 1.01 | 0.98-1.05 | 0.99 | 0.95-1.03 | 1.00 | 0.98-1.02 | 1.01 | 1-1.02 | 1.00 | 0.99-1.01 | 0.99 | 0.97-1.01 |
| -1 | 1.00 | 0.98-1.03 | 1.00 | 0.98-1.01 | 1.00 | 0.98-1.03 | 1.00 | 0.96-1.05 | 1.00 | 0.98-1.02 | 1.00 | 0.99-1.01 | 1.00 | 0.99-1.01 | 1.00 | 0.99-1.02 |
| 0 | 0.99 | 0.97-1.02 | 0.97 | 0.94-0.99 | 1.00 | 0.98-1.02 | 1.00 | 0.98-1.02 | 1.00 | 0.99-1.01 | 0.99 | 0.98-1 | 1.01 | 1-1.02 | 0.95 | 0.93-0.97 |
| 1 | 0.89 | 0.84-0.95 | 0.87 | 0.81-0.92 | 0.96 | 0.91-1.02 | 0.96 | 0.89-1.02 | 0.99 | 0.92-1.06 | 0.99 | 0.97-1.02 | 1.01 | 0.98-1.04 | 0.68 | 0.61-0.76 |
| 2 | 0.79 | 0.72-0.86 | 0.82 | 0.75-0.89 | 0.94 | 0.86-1.03 | 0.87 | 0.78-0.96 | 0.97 | 0.87-1.08 | 0.97 | 0.93-1.01 | 0.97 | 0.93-1.01 | 0.55 | 0.47-0.64 |
| 3 | 0.64 | 0.55-0.73 | 0.78 | 0.7-0.86 | 0.95 | 0.85-1.06 | 0.81 | 0.7-0.94 | 0.91 | 0.8-1.03 | 0.95 | 0.91-1 | 0.94 | 0.89-0.99 | 0.47 | 0.38-0.57 |
| 4 | 0.55 | 0.45-0.67 | 0.76 | 0.66-0.87 | 0.91 | 0.78-1.06 | 0.77 | 0.64-0.92 | 0.85 | 0.73-0.99 | 0.95 | 0.88-1.01 | 0.89 | 0.83-0.95 | 0.41 | 0.33-0.52 |
| 5 | 0.52 | 0.41-0.67 | 0.74 | 0.64-0.85 | 0.86 | 0.72-1.03 | 0.73 | 0.58-0.92 | 0.82 | 0.68-0.98 | 0.89 | 0.82-0.97 | 0.85 | 0.78-0.92 | 0.33 | 0.25-0.44 |
| 6 | 0.53 | 0.41-0.67 | 0.72 | 0.62-0.84 | 0.79 | 0.63-0.98 | 0.66 | 0.52-0.86 | 0.71 | 0.57-0.87 | 0.86 | 0.77-0.96 | 0.82 | 0.73-0.91 | 0.28 | 0.2-0.38 |
| 7 | 0.45 | 0.34-0.6 | 0.69 | 0.59-0.83 | 0.82 | 0.65-1.04 | 0.63 | 0.46-0.85 | 0.61 | 0.46-0.81 | 0.84 | 0.75-0.94 | 0.81 | 0.73-0.91 | 0.22 | 0.15-0.34 |
| 8 | 0.36 | 0.24-0.54 | 0.73 | 0.59-0.89 | 0.70 | 0.54-0.91 | 0.52 | 0.35-0.76 | 0.54 | 0.37-0.78 | 0.82 | 0.7-0.96 | 0.68 | 0.58-0.79 | 0.18 | 0.11-0.29 |

Notes: Control group: never treated, 0 anticipation periods, doubly robust estimation, Callaway-Sant'Anna staggered difference-in-differences method.

### 3.3 Competition

Overall, we found that after patent expiration the increase in the average number of generic competitors entering the market differed strongly between countries. Figure S2 presents the event study estimates.

We found the highest number of competitors in Germany, with approximately 15 competitors entering the market up to eight years after patent expiration, followed by Japan (∼10 competitors), France (∼7 competitors), and the US (∼7 competitors). Conversely, Australia, Canada, Switzerland, and the UK presented the lowest number of competitors with less than 5 generics entering the market after patent expiration. Figure S3 presents all the point estimates to ease interpretation.

When examining the influence on the price per competitor, we found the greatest effect size for the US, Figure S4 (standardized coefficients by two standard deviations). The estimates suggest that price competition varied greatly, and seemed to be independent from the number of competitors that entered the market. Figure S5 presents the estimates for both linear and quadratic coefficients of amount of competition. Overall, the estimates of the linear terms were stable.

### 3.3 Cost-effectiveness effects

The main results of our cost-effectiveness simulation models are presented in Figure 2 and Figure 3. All the simulations incorporated the US genericization pricing estimates.

**Figure 2.**
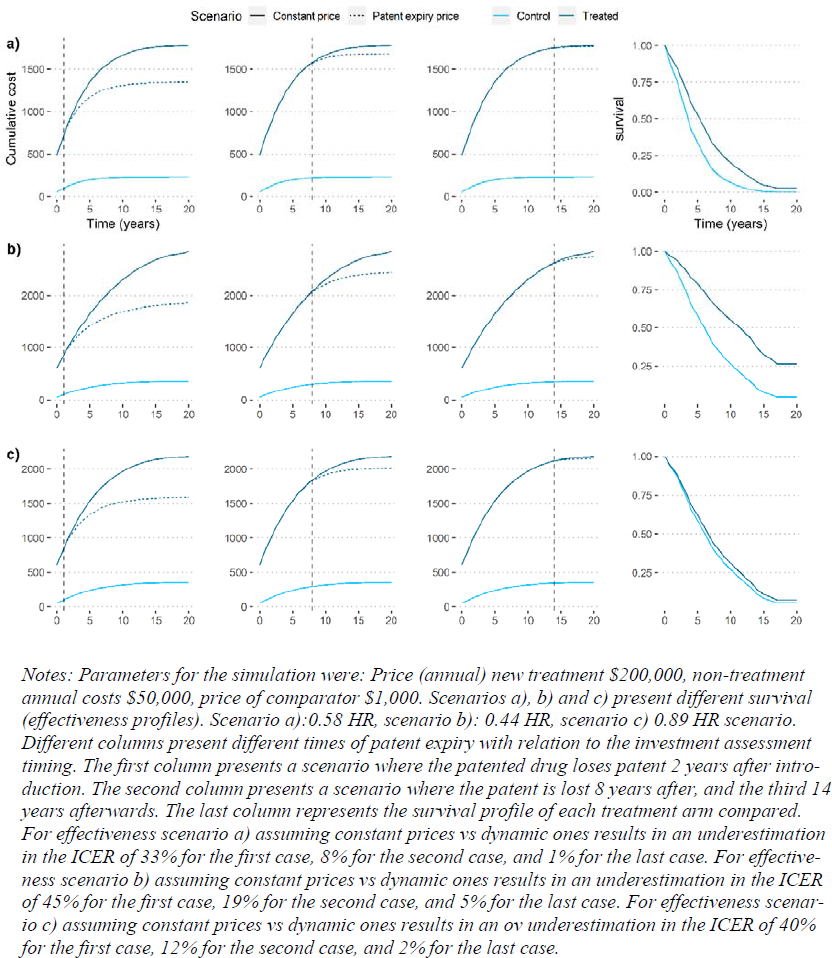
Cost-effectiveness simulation scenarios with and without post-patent expiry prices for the new originator drug.

**Figure 3.**
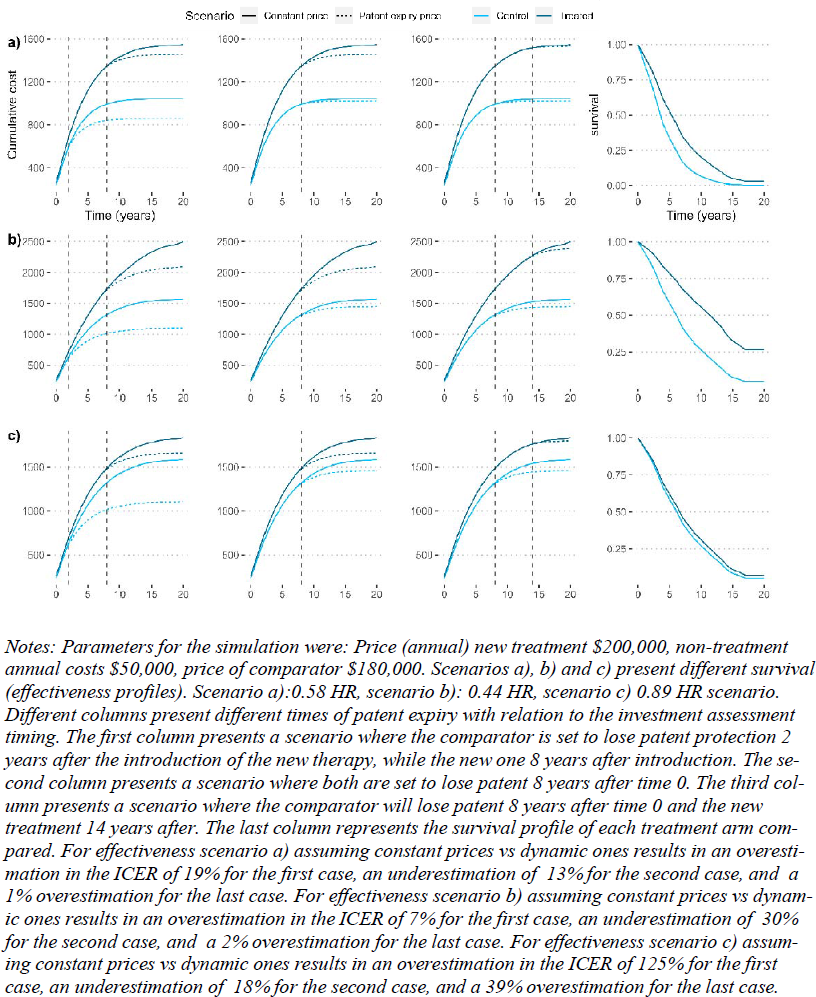
Cost-effectiveness simulation scenarios with and without post-patent expiry prices for the new originator drugs and their control arms.

In the first scenario, we compared two drugs indicated for the same treatment. The first drug was a new originator drug with an annual price of $200,000, the second drug was a generic comparator with an annual price of $1,000, and annual non-pharmaceutical costs of $50,000, over a 20-year time horizon. The model only included overall survival as the outcome of effectiveness, differentiating between three subscenarios – moderate, high, and low effectiveness. We then analyzed the ICERs for different scenarios, where patent expiration occurred two, eight and 14 years after the assessment. Our model revealed that if patent expiration occurs 14 years after market entry, ignoring price dynamics after patent expiration has a very small impact on ICERs, with an underestimation of 1%, 2%, and 5%, for the low, mid and high effectiveness cases. In contrast, if patent expiry occurs one year after the ICER assessment, the underestimation ranges from 33% to 40% of the ICER with constant prices.

In the second scenario, we compared two drugs indicated for the same treatment, both under patent protection at the time of ICER assessment. The new originator drug had an annual price of $200,000. The comparator drug had an annual price of $180,000, and annual non-pharmaceutical costs of $50,000, with the same time horizon, effectiveness and discount rates as in the first scenario. We analysed three subscenarios: In the first subscenario, the comparator drug lost its patent two years after time zero and the new originator drug eight years after. In the second subscenario, the patent of both drugs expired eight years after time zero. In the final subscenario, the patent of the comparator drug expired eight years after time zero, while the patent for the new originator drug expired 14 years after time zero. We found that ignoring post-patent pricing dynamics had a significant impact on es-timated ICERs, with an ICER overestimation ranging from 19 to 125% when the comparator’s patent expired prior to the new originator drug.

The results can be summarized as follows: First, if the new originator drug under evaluation has a high effectiveness profile resulting in prolonged therapy administration (i.e., a drug for a chronic illness that is administered to the patient until death or progression), ignoring post-patent price dynamics underestimates the true ICER in cases where the comparator is a generic drug. Second, if the comparator drug is still under patent protection at the time of evaluation, but the patent will expire prior to the patent of the new originator drug, ignoring pricing dynamics severely overestimates its cost-effectiveness. Third, the closer the time duration between CEA and patent expiration, the higher is the bias of the true ICER.

## 4. Discussion

In this study, we present credible estimates of drug pricing dynamics and on the effects of patent protection loss on drug prices for eight developed economies. Our estimates demonstrate a generalized price decrease across nations after market entry of generic competitors, with a high degree of heterogeneity across countries. Stark differences in price trajectories before patent loss across countries were observed, likely reflecting the different priority settings and price negotiations for each healthcare system (Vokinger et al., 2021).

Our estimates revealed that the strongest price declines after patent expiration occurred in the US. However, each included country in our study presented its own particular dynamics. Our estimates can be incorporated into cost-effectiveness analyses, either as sensitivity analyses or for base-case scenarios (Woods et al., 2021). Additionally, our estimates can also be applied to models where both comparators are under patent protection at the time of the cost-effectiveness analysis, but the patent of the comparator drug will expire earlier than the new originator drug. A price decline of the comparator drug with a constant price of the new originator drug results in an overestimation of the cost-effectiveness of the new originator dug.

Our study has limitations. First, our dataset contained list prices for several countries. Many countries have initiated the incorporation of (confidential) rebates, for which we could not account for. If such rebates increase after patent loss, our estimates are biased upwards. Second, the comparability of prices across countries is compromised due to the difference in data sources extracted by IMS data.

## 5. Conclusion

We estimated significant decline in drug prices after patent expiration, ranging from 30 to 80% depending on the country eight years after patent expiration. Our results further showed that if the new originator drug under evaluation has a high effectiveness profile resulting in prolonged therapy administration, ignoring post-patent price dynamics underestimates the true ICER in cases where the comparator is a generic drug. Furthermore, if the comparator drug is still under patent protection at the time of evaluation, but the patent will expire prior to the patent of the new originator drug, ignoring pricing dynamics severely overestimates its cost-effectiveness. Lastly, the closer the time duration between CEA and patent expiration, the higher is the bias of the true ICER.

Our pricing dynamic estimates can be applied to base-case analyses of cost-effectiveness models across the lifecycle of drugs in the US, Australia, Canada, France, Germany, Japan, Switzer-land, and the UK.

## Data Availability

Data cannot be shared due to being property of IQVIA.

## Supplementary Material

### Supplementary Cost-effectiveness model summary

Assume that the discount rates for both costs and effectiveness are the same (r_e_=r_c_) and further assume that the non-pharmaceutical cost of the treatment and the control group are equal (c_bt_=c_at_).

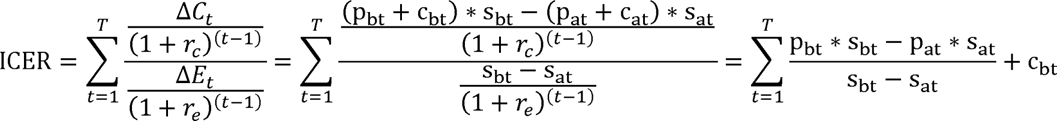

In the last step the tow assumptions were used namely that 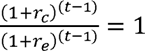 and that

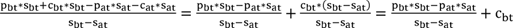

Further assuming that c_bt_ and p_at_ are constant over time (c_b_, p_a_) we can simplify to:

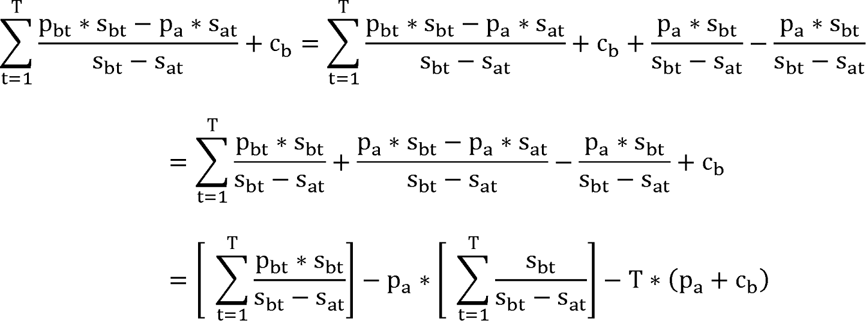

Using in the last step that we can some over p_a_ and c_b_ as they are constant in time.

Finally, by recentering the observations we can set p_a_ and c_b_ to 0, meaning that the control group has assumed cost zero, and then derive the final expression for ICER:

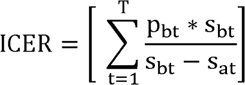

## Supplementary Results

**Figure S1.1.**
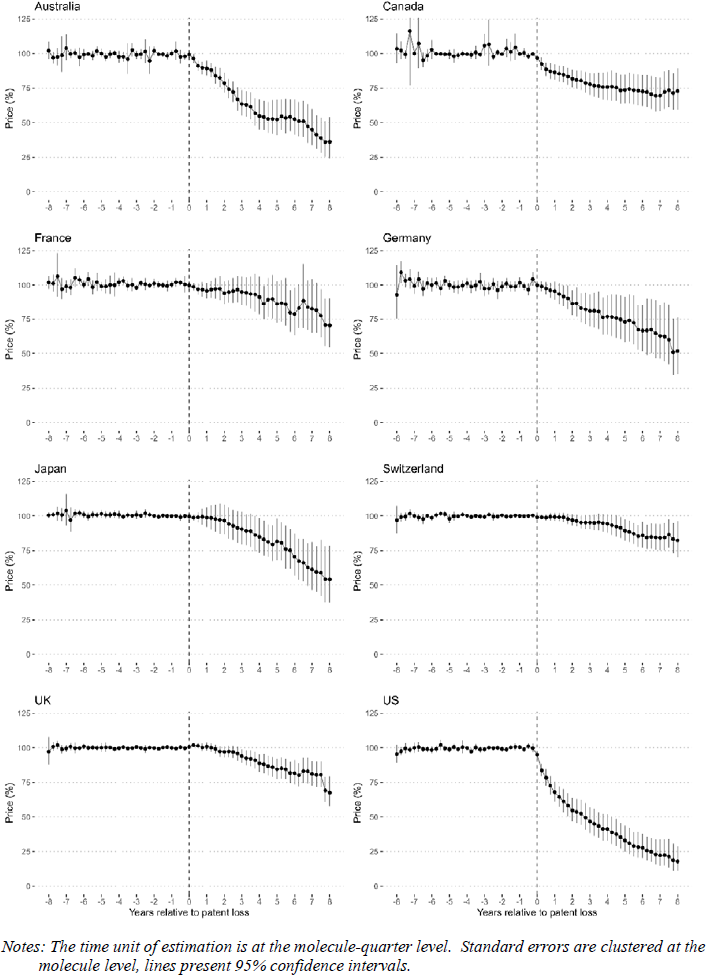
Event study estimates of patent loss and prices, Callaway-Sant’Anna.

**Figure S1.2.**
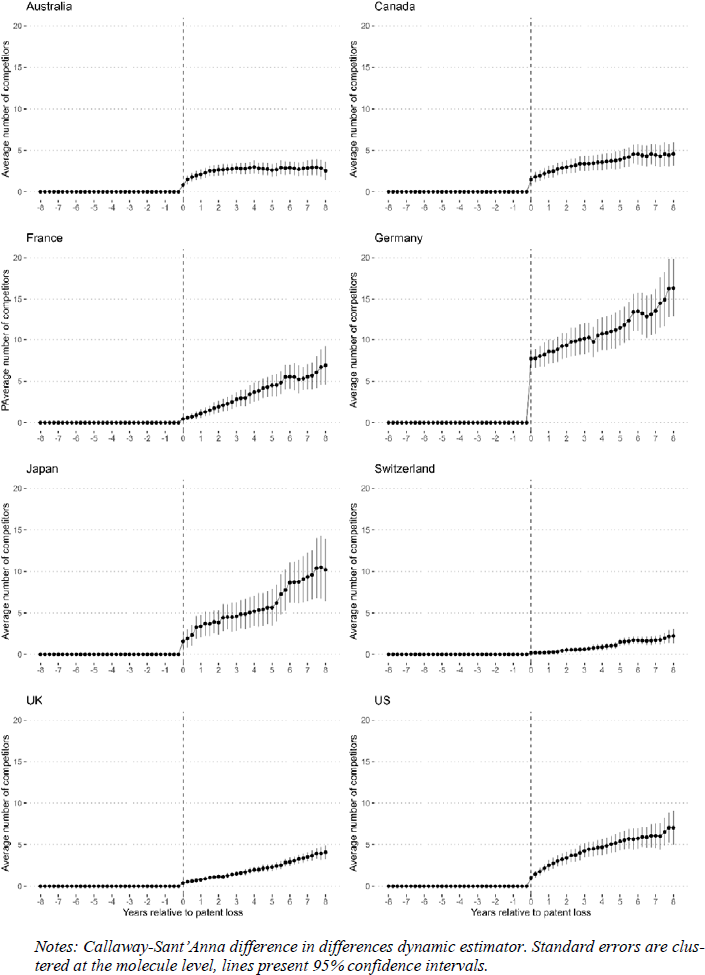
Event study estimates of patent loss and amount of competitors in the market.

**Figure S1.3.**
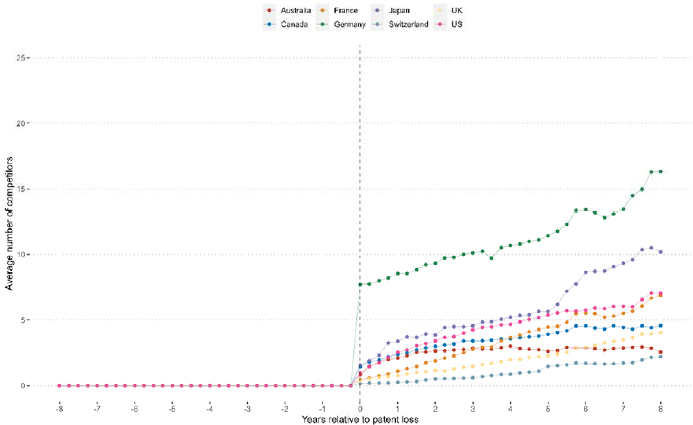
Event study estimates of patent expiration and number of competitors entering the market after patent expiration.

**Figure S1.4.**
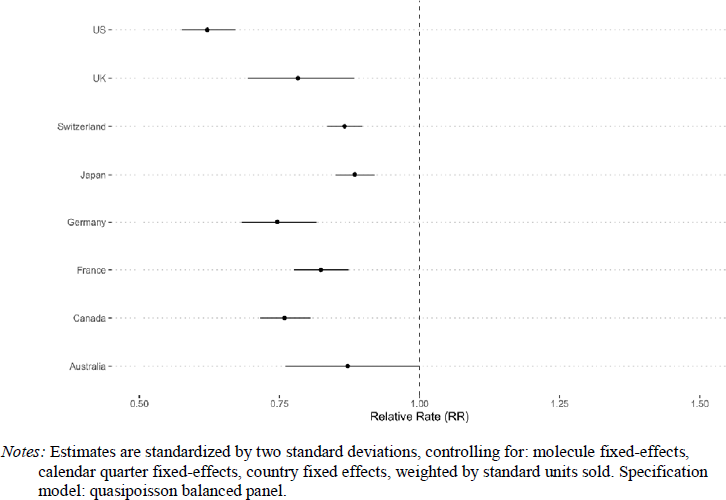
Estimates on the marginal effect of one additional competitor drug on prices of new originator drugs.

**Figure S1.5.**
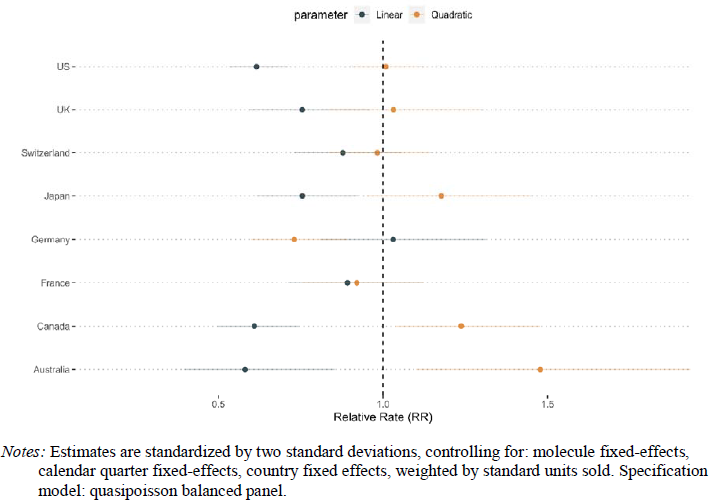
Estimates on the effects of additional competitors drug on prices of new originator drugs, linear and quadratic effects.

**Table S1.** Distribution of the drugs in the study cohort classified in therapeutic areas by countries (ATC-1^st^-level) and metabolism forming organs system systemic use system tem

| **ATC group code** | **Australia** | **Canada** | **France** | **Germany** | **Japan** | **Switzerland** | **UK** | **US** |
| --- | --- | --- | --- | --- | --- | --- | --- | --- |
| Alimentary tract and metabolism | 46 | 37 | 26 | 35 | 52 | 30 | 35 | 56 |
| Blood and blood forming organs | 18 | 21 | 18 | 26 | 15 | 19 | 25 | 39 |
| Cardiovascular system | 21 | 35 | 25 | 35 | 31 | 38 | 33 | 40 |
| Dermatologicals | 9 | 10 | 4 | 8 | 9 | 5 | 7 | 11 |
| Genito urinary system and sex hormones | 12 | 8 | 11 | 9 | 7 | 10 | 10 | 15 |
| Systemic hormonal preparations, excl. sex hormones and insulins | 4 | 7 | 6 | 9 | 5 | 4 | 5 | 8 |
| Anti-infective for systemic use | 57 | 68 | 69 | 70 | 62 | 61 | 65 | 83 |
| Antineoplastic and immunomodulating agents | 88 | 94 | 96 | 106 | 97 | 86 | 97 | 118 |
| Musculo-skeletal system | 15 | 10 | 11 | 10 | 11 | 14 | 15 | 18 |
| Nervous system | 36 | 36 | 25 | 32 | 31 | 32 | 36 | 63 |
| Respiratory system | 19 | 18 | 16 | 20 | 17 | 14 | 18 | 21 |
| Sensory organs | 8 | 15 | 7 | 9 | 12 | 7 | 8 | 12 |
| Various | 2 | 2 | 3 | 6 | 3 | 1 | 2 | 4 |

**Table S2.**
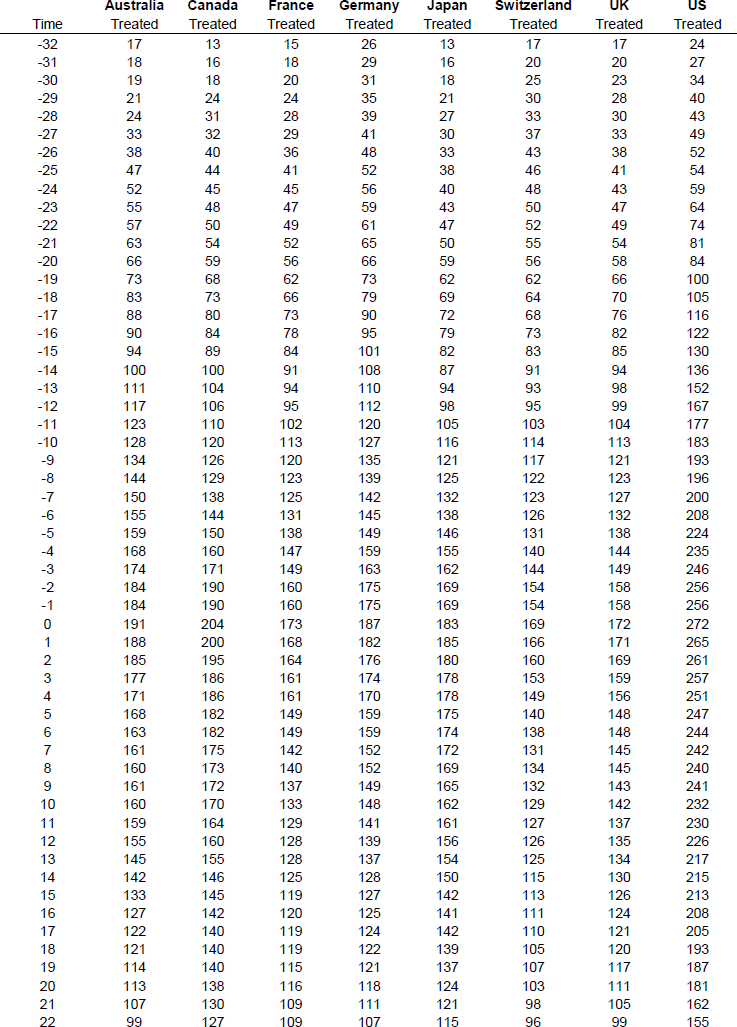
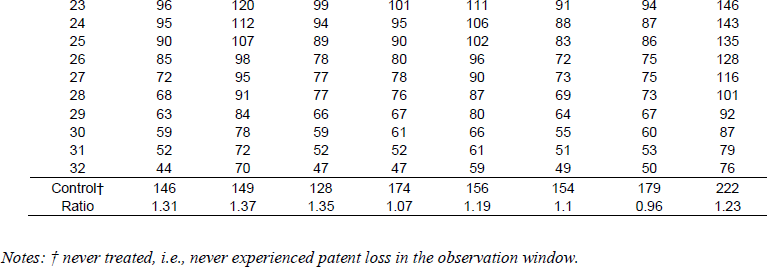
Time distribution of observation with and without patent loss by country.

| Time | Australia<br>Treated | Canada<br>Treated | France<br>Treated | Germany<br>Treated | Japan<br>Treated | Switzerland<br>Treated | UK<br>Treated | US<br>Treated |
| --- | --- | --- | --- | --- | --- | --- | --- | --- |
| -32 | 17 | 13 | 15 | 26 | 13 | 17 | 17 | 24 |
| -31 | 18 | 16 | 18 | 29 | 16 | 20 | 20 | 27 |
| -30 | 19 | 18 | 20 | 31 | 18 | 25 | 23 | 34 |
| -29 | 21 | 24 | 24 | 35 | 21 | 30 | 28 | 40 |
| -28 | 24 | 31 | 28 | 39 | 27 | 33 | 30 | 43 |
| -27 | 33 | 32 | 29 | 41 | 30 | 37 | 33 | 49 |
| -26 | 38 | 40 | 36 | 48 | 33 | 43 | 38 | 52 |
| -25 | 47 | 44 | 41 | 52 | 38 | 46 | 41 | 54 |
| -24 | 52 | 45 | 45 | 56 | 40 | 48 | 43 | 59 |
| -23 | 55 | 48 | 47 | 59 | 43 | 50 | 47 | 64 |
| -22 | 57 | 50 | 49 | 61 | 47 | 52 | 49 | 74 |
| -21 | 63 | 54 | 52 | 65 | 50 | 55 | 54 | 81 |
| -20 | 66 | 59 | 56 | 66 | 59 | 56 | 58 | 84 |
| -19 | 73 | 68 | 62 | 73 | 62 | 62 | 66 | 100 |
| -18 | 83 | 73 | 66 | 79 | 69 | 64 | 70 | 105 |
| -17 | 88 | 80 | 73 | 90 | 72 | 68 | 76 | 116 |
| -16 | 90 | 84 | 78 | 95 | 79 | 73 | 82 | 122 |
| -15 | 94 | 89 | 84 | 101 | 82 | 83 | 85 | 130 |
| -14 | 100 | 100 | 91 | 108 | 87 | 91 | 94 | 136 |
| -13 | 111 | 104 | 94 | 110 | 94 | 93 | 98 | 152 |
| -12 | 117 | 106 | 95 | 112 | 98 | 95 | 99 | 167 |
| -11 | 123 | 110 | 102 | 120 | 105 | 103 | 104 | 177 |
| -10 | 128 | 120 | 113 | 127 | 116 | 114 | 113 | 183 |
| -9 | 134 | 126 | 120 | 135 | 121 | 117 | 121 | 193 |
| -8 | 144 | 129 | 123 | 139 | 125 | 122 | 123 | 196 |
| -7 | 150 | 138 | 125 | 142 | 132 | 123 | 127 | 200 |
| -6 | 155 | 144 | 131 | 145 | 138 | 126 | 132 | 208 |
| -5 | 159 | 150 | 138 | 149 | 146 | 131 | 138 | 224 |
| -4 | 168 | 160 | 147 | 159 | 155 | 140 | 144 | 235 |
| -3 | 174 | 171 | 149 | 163 | 162 | 144 | 149 | 246 |
| -2 | 184 | 190 | 160 | 175 | 169 | 154 | 158 | 256 |
| -1 | 184 | 190 | 160 | 175 | 169 | 154 | 158 | 256 |
| 0 | 191 | 204 | 173 | 187 | 183 | 169 | 172 | 272 |
| 1 | 188 | 200 | 168 | 182 | 185 | 166 | 171 | 265 |
| 2 | 185 | 195 | 164 | 176 | 180 | 160 | 169 | 261 |
| 3 | 177 | 186 | 161 | 174 | 178 | 153 | 159 | 257 |
| 4 | 171 | 186 | 161 | 170 | 178 | 149 | 156 | 251 |
| 5 | 168 | 182 | 149 | 159 | 175 | 140 | 148 | 247 |
| 6 | 163 | 182 | 149 | 159 | 174 | 138 | 148 | 244 |
| 7 | 161 | 175 | 142 | 152 | 172 | 131 | 145 | 242 |
| 8 | 160 | 173 | 140 | 152 | 169 | 134 | 145 | 240 |
| 9 | 161 | 172 | 137 | 149 | 165 | 132 | 143 | 241 |
| 10 | 160 | 170 | 133 | 148 | 162 | 129 | 142 | 232 |
| 11 | 159 | 164 | 129 | 141 | 161 | 127 | 137 | 230 |
| 12 | 155 | 160 | 128 | 139 | 156 | 126 | 135 | 226 |
| 13 | 145 | 155 | 128 | 137 | 154 | 125 | 134 | 217 |
| 14 | 142 | 146 | 125 | 128 | 150 | 115 | 130 | 215 |
| 15 | 133 | 145 | 119 | 127 | 142 | 113 | 126 | 213 |
| 16 | 127 | 142 | 120 | 125 | 141 | 111 | 124 | 208 |
| 17 | 122 | 140 | 119 | 124 | 142 | 110 | 121 | 205 |
| 18 | 121 | 140 | 119 | 122 | 139 | 105 | 120 | 193 |
| 19 | 114 | 140 | 115 | 121 | 137 | 107 | 117 | 187 |
| 20 | 113 | 138 | 116 | 118 | 124 | 103 | 111 | 181 |
| 21 | 107 | 130 | 109 | 111 | 121 | 98 | 105 | 162 |
| 22 | 99 | 127 | 109 | 107 | 115 | 96 | 99 | 155 |
| 23 | 96 | 120 | 99 | 101 | 111 | 91 | 94 | 146 |
| 24 | 95 | 112 | 94 | 95 | 106 | 88 | 87 | 143 |
| 25 | 90 | 107 | 89 | 90 | 102 | 83 | 86 | 135 |
| 26 | 85 | 98 | 78 | 80 | 96 | 72 | 75 | 128 |
| 27 | 72 | 95 | 77 | 78 | 90 | 73 | 75 | 116 |
| 28 | 68 | 91 | 77 | 76 | 87 | 69 | 73 | 101 |
| 29 | 63 | 84 | 66 | 67 | 80 | 64 | 67 | 92 |
| 30 | 59 | 78 | 59 | 61 | 66 | 55 | 60 | 87 |
| 31 | 52 | 72 | 52 | 52 | 61 | 51 | 53 | 79 |
| 32 | 44 | 70 | 47 | 47 | 59 | 49 | 50 | 76 |
| Control† | 146 | 149 | 128 | 174 | 156 | 154 | 179 | 222 |
| Ratio | 1.31 | 1.37 | 1.35 | 1.07 | 1.19 | 1.1 | 0.96 | 1.23 |

Notes: † never treated, i.e., never experienced patent loss in the observation window.

